# GC-MS Analysis of Smokeless tobacco (Shamma): Toxicological Evaluation

**DOI:** 10.1101/2020.04.23.20072553

**Authors:** Hamzah S. Alzahrani, Tariq S. Alamoudi, Khaled S. Aloudah, Fahad S. Alghamdi, Khalel S. Alghamdi, Haytham A. Ali

## Abstract

**Background:** Smoking is a badly addictive habit due to nicotine contents in tobacco. Shammah is one way of smokeless tobacco that is homely made and it use resulting in multiple medical issues.

**Aim:** The aim of the present study were to chemically analyze the content of various types of Shammah using GC-MS analysis with clinical biochemical investigation of some cases usually used shamma in Makkah region, Saudi Arabia.

**Methods:** Three samples from two main different types of *shamma (black and yellow)* were collected from local providers and analyzed by Gas Chromatography Mass Spectrometry (GC/MS). Eighteen blood samples were also collected from apparently healthy male peoples (30-45 years) classified into 3 groups; group 1 (control) people who doesn’t smoke or use any type of shamma, second and third groups were people who usually used black and yellow shamma respectively; CBC and biochemical analysis were performed.

**Results:** In GC-MS Analysis, the nicotine represents the major constituent in all samples. Total RBCs count, Hemoglobin (HGB) Conc., eosinophil % and iron, total cholesterol, HDL-c concentrations were significantly decreased in peoples using either black or yellow shamma whereas serum levels of ALT, AST and ALP enzyme activities, BUN and creatinine levels were significantly increases.

**Conclusion:** Different shamma samples contains different concentrations of various toxic compounds that can produced a serious health problems as hypochromic anemia due to iron deficiency, decrease in eosinophil %, with hepatic and renal cell injuries detected as increase ALT, AST, ALP, BUN, and creatinine levels.

## Introduction

Smoking is considered as epidemic as more than one billion addicting it due to presence of nicotine in tobacco mostly in developing countries and in males more than females **[1**,**2]**. Although tobacco is legally sold and regulated by FDA **[3]** but it has a high mortality number over eight million every year according to World Health Organization (WHO) **[1]**. Two main types of tobacco has been identified; usual smoking and smokeless tobacco (ST) **[4]**. ST has become a worldwide medical problem with around 350 million clients, widely used in the United States, Sweden, Norway and Asia; the highly number of user are women and young people. It is utilized by mouth via chewing or holding between the cheek and gum with very high addiction due to its nicotine contents **[5, 6]**.

In Saudi Arabian population, the maximum use of smokeless tobacco was observed in southern region near to Yemen and overspread over all the Kingdome. There are many common smokeless tobacco products in the eastern Mediterranean region such as betel quid with tobacco, Naswar, Toombak. Dry snuff and Shamma **[7]**.

Locally, *Shamma* is manufactured form of smokeless tobacco, the addictive substance which is used by placing in the oral cavity, producing saliva that then ingested. It is generally mainly produced from tobacco leaves such as *N. glauca, N. tabacum, N. nepalensis or N*.*rustica* **[8**,**9]**. It was reported that, Shamma contains at least 28 of potential carcinogenic substance with nitrosamines as the most harmful **[10]**. Shamma is a mixture of ground tobacco leafs, lime, ash, and flavorings. Nicotine is the main ingredient in shamma and accounts for 98% of total alkaloids contained in tobacco leaf in some types of *nicotiana species*. Moreover various unsafe substances, including 28 potential cancer-causing agents have been identified in Shammah among these substances nitrosamines are accounted for to be the most destructive **[11]**. Shamma is an example of custom-made smokeless tobacco which is prepared at home and is consumed shortly after preparation **[12]**.

Other reports have gone even further and assumed that the TSNAs (i.e., NNAL) that exist in shamma products are lung cancer-specific carcinogens **[13]**.

Despite the fact that nicotine itself is not carcinogenic, its derivatives such as 4-(methylnitrosamino)-1-(3-pyridyl)-1-butanone and *N*′-nitrosonornicotine might be responsible for the carcinogenic effect **[14]**.

Moderate and reversible toxicity along with weight loss was found in the esophagus, stomach, liver, kidneys, and lungs **[15]**. (Presence of ammonia, benzo[a]pyrene, cadmium, nickel, nicotine, nitrates, and tobacco-specific nitrosamines increase the risk of probabilistic cancer **[16]**. Heavy metals such as arsenic and nickel have synergistic effects with risk factors associated with oral cancer **[17]**.

Shammah also have been reported to promote RBC membrane damage and increased inflammation in systematic stress **[18]**. Thus, more studies are needed to provide evidence-based data to health authorities and Ministry of Interior to enhance in the design of sound strategies to thwart it. In the light of the above mentioned, our study aimed to chemically analyze the content of different samples of shamma types using GC-MS analysis with a clinical biochemical investigation of blood samples collected from different apparently healthy people either used black or yellow shamma or not smokers at all.

## Material and methods

### Chemicals and reagents

a. Methanol CH3OH needed for sample preparation were supplied by Panreac.
b. A nicotine standard (purity 99.0%, batch No. S18379725) was purchased from Toronto-Research-Chemicals, Inc. (TRC, Toronto, ON, Canada).
c. An internal standard, fennel (purity 99.0%, batch No. ED4XO-ON) was purchased from J & K Chemical Ltd. (Shanghai, China).
d. Mass spectroscopy grade acetonitrile, methanol, and formic acid were also purchased from J & K Chemical Ltd. (Shanghai, China).

### Selection of samples and GC-MS analysis

Six different shamma samples were purchased from markets, 3 from each main types black and yellow shamma. S1, S2 and S3 were samples from what’s called *Black shamma whereas S4, S5 and S6 were samples* from Arishi *shamma* (yellow in color and is also called as yellow *shamma*).

Shamma powder was stored at 4 □C, protected from light and moisture till GC-MS analysis using Gas Chromatography Mass Spectrometry (GC/MS) 5973 inert Mass selective Detector 6890N Network GC system from Agilent Technologies which separates different compounds in the sample into pulses of pure chemicals based on their volatility by flowing an inert gas (mobile phase), which carries the sample through a (stationary phase) fixed in the column. Spectra of compounds are collected as they exit a chromatographic column by the mass spectrometer, which quantifies and identifies the chemicals according to their mass to charge ratio (m/z). These spectra then are stored on the computer and analyzed **[19]**.

### Cases and experimental design

Eighteen apparently healthy males (30-45 year) were volunteered for this experiment divided into 3 groups (6 persons each), first considered as a control group who didn’t smoke or receive any type of shamma, second were the persons used to use black shamma while the third were the persons used to use yellow shamma. Blood samples were collected from 18 persons in vacuum tube containers with ethylene-diamine-tetra-acetic acid (EDTA). The experiments were conducted after ethical approval by the local authorities at University of Jeddah, Saudi Arabia and after personal agreement of all volunteers. Hematological examination was performed using a Bayer ADVIA2120 automatic blood cell analyzer (Leverkusen, Germany) and a MERLIN MCLOplus (Leverkusen, Germany) to measure CBC parameters. Blood for clinical chemistry was collected in vacuum tubes devoid of anticoagulant, allowed to clot at room temperature, centrifuged, and then serum was separated. The serum biochemical parameters were assessed using a HITACHI 7080 automated biochemical analyzer (Tokyo, Japan) and an Easylyte PLUS electrolyte analyzer (MEDICA, Bedford, MA, America).

### Statistical Analysis

All measurements are expressed as the mean ± standard error. For all samples hematological parameters and clinical chemistry data were analyzed by parametric one-way analysis using the F-test (ANOVA) with Statistical Product and Service Solutions (SPSS) v22 (IBM: Armonk, America). If the resulting p-value was <0.05, a comparison of each group using the LSD test was performed for the hypothesis of equal means. The Dunnett T3 test was applied when the data could not be assumed to follow homogeneity variance.

## Results

**Figure 1:**
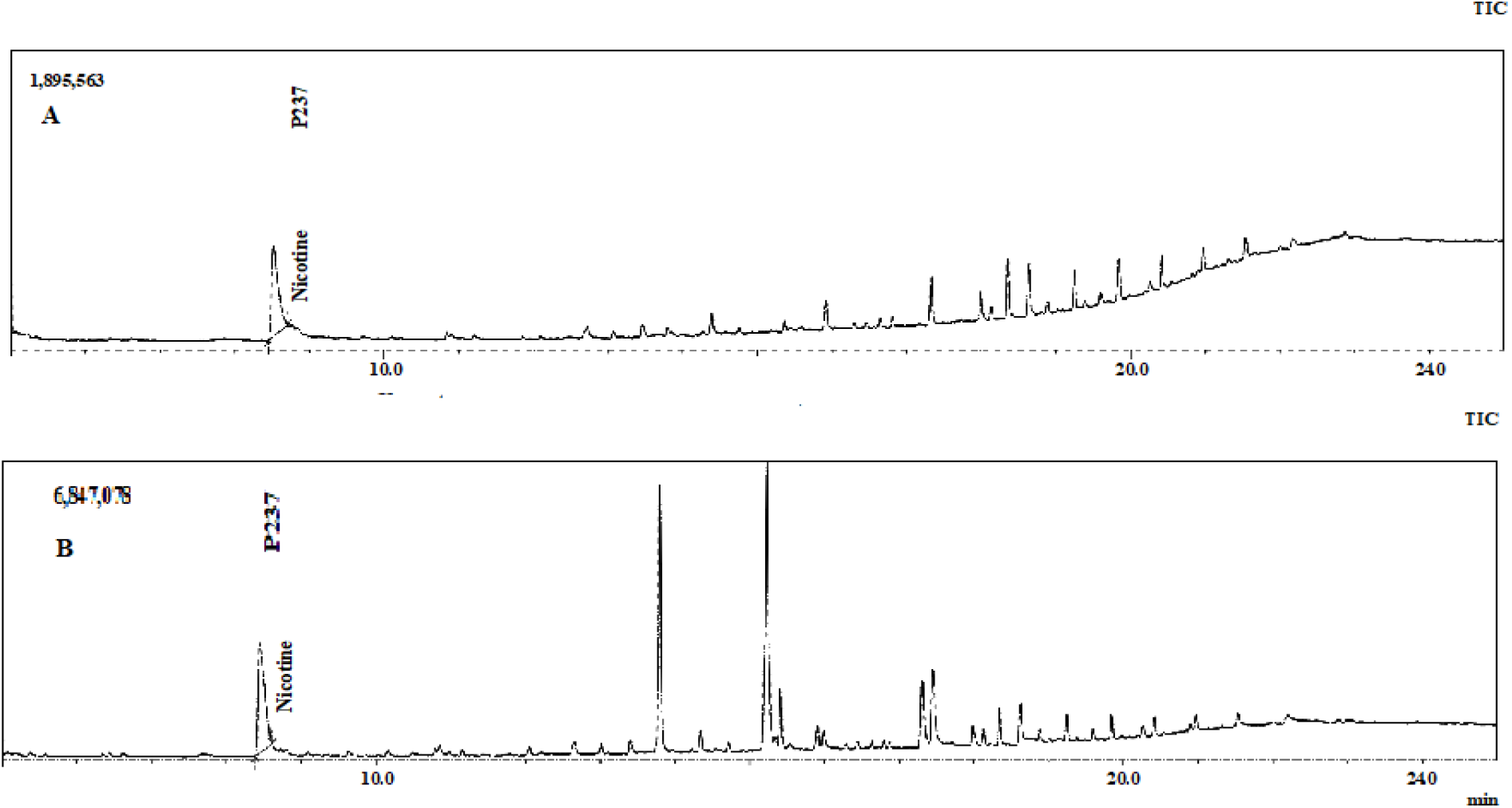
Chromatogram test and peak report of black shamma (A) and yellow shamma (B) Using GC-MS analysis.

Representative chromatograms showed the separated and identified constituents of black and yellow *shamma* samples efficiently. Whereas all components identified in *shamma* samples are listed in Table 1.

**Table 1:**
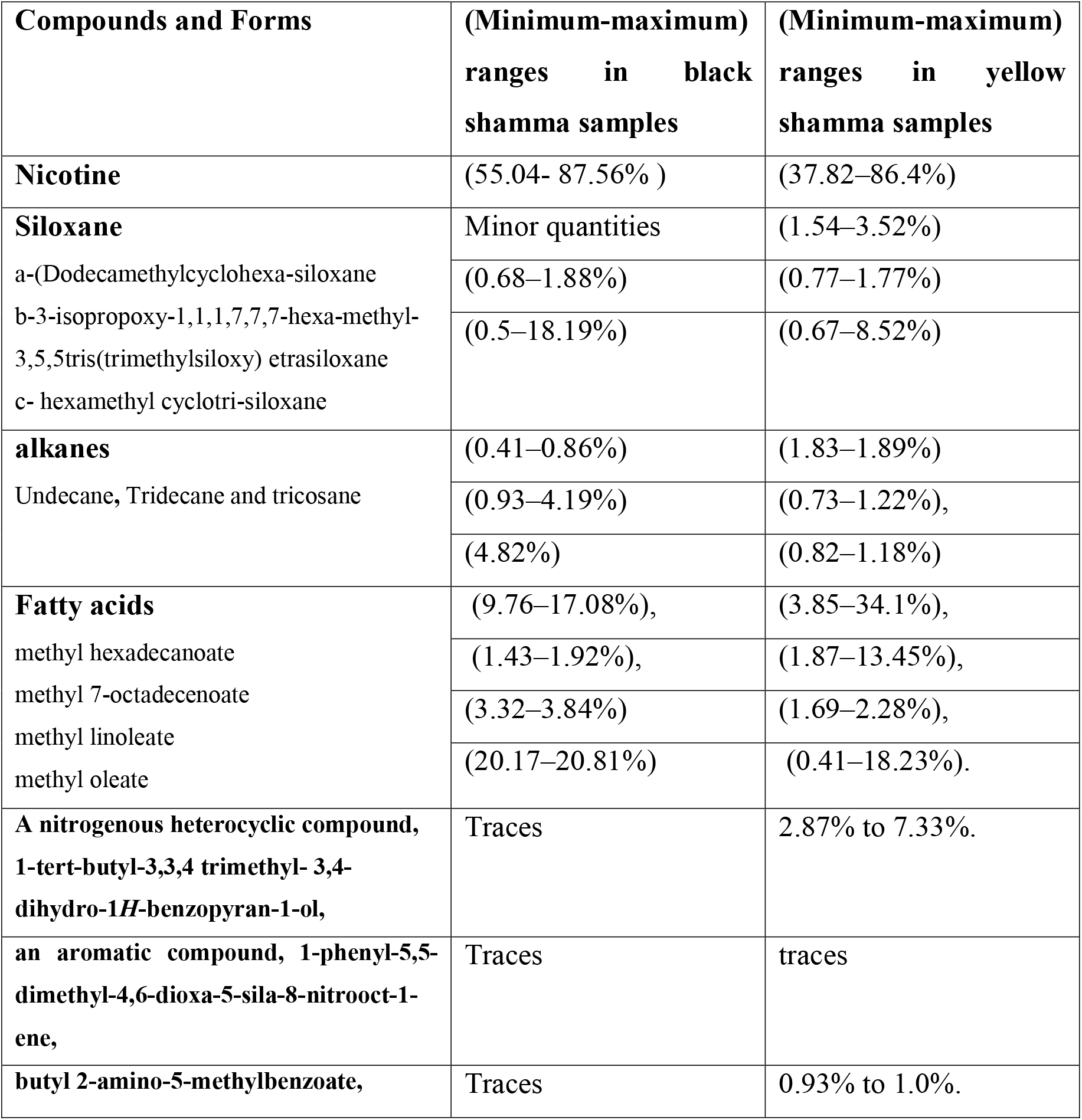
Major constituents, forms and their concentrations in two main shamma samples.

| **Compounds and Forms** | **(Minimum-maximum) ranges in black shamma samples** | **(Minimum-maximum) ranges in yellow shamma samples** |
| --- | --- | --- |
| **Nicotine** | (55.04- 87.56% ) | (37.82–86.4%) |
| **Siloxane** | Minor quantities | (1.54–3.52%) |
| a-(Dodecamethylcyclohexa-siloxane | (0.68–1.88%) | (0.77–1.77%) |
| b-3-isopropoxy-1,1,1,7,7,7-hexa-methyl-3,5,5tris(trimethylsiloxy) etrasiloxane | (0.5–18.19%) | (0.67–8.52%) |
| c- hexamethyl cyclotri-siloxane |  |  |
| **alkanes** | (0.41–0.86%) | (1.83–1.89%) |
| Undecane, Tridecane and tricosane | (0.93–4.19%) | (0.73–1.22%), |
|  | (4.82%) | (0.82–1.18%) |
| **Fatty acids** | (9.76–17.08%), | (3.85–34.1%), |
| methyl hexadecanoate | (1.43–1.92%), | (1.87–13.45%), |
| methyl 7-octadecenoate | (3.32–3.84%) | (1.69–2.28%), |
| methyl linoleate | (20.17–20.81%) | (0.41–18.23%). |
| methyl oleate |  |  |
| **A nitrogenous heterocyclic compound, 1-tert-butyl-3,3,4 trimethyl- 3,4-dihydro-1H-benzopyran-1-ol,** | Traces | 2.87% to 7.33%. |
| **an aromatic compound, 1-phenyl-5,5-dimethyl-4,6-dioxa-5-sila-8-nitrooct-1-ene,** | Traces | traces |
| **butyl 2-amino-5-methylbenzoate,** | Traces | 0.93% to 1.0%. |

The GC-MS analysis of all the samples revealed the presence of similar groups of constituents including alkanes, siloxanes, pyridine derivatives, fatty acid esters, and amides (Table 1).

Nicotine was extracted mainly in the nonpolar solvents, and it represents the major constituent in all samples.

The presented results in table 2 showed that, there was a significant decrease in the total RBCs count, HGB concentration, eosinophil and RDW-SD in peoples usually use any types of shamma which suggests hypochromic anemia and eosinopenia with a decrease in the width of the red cell distribution curve.

**Table 2:**
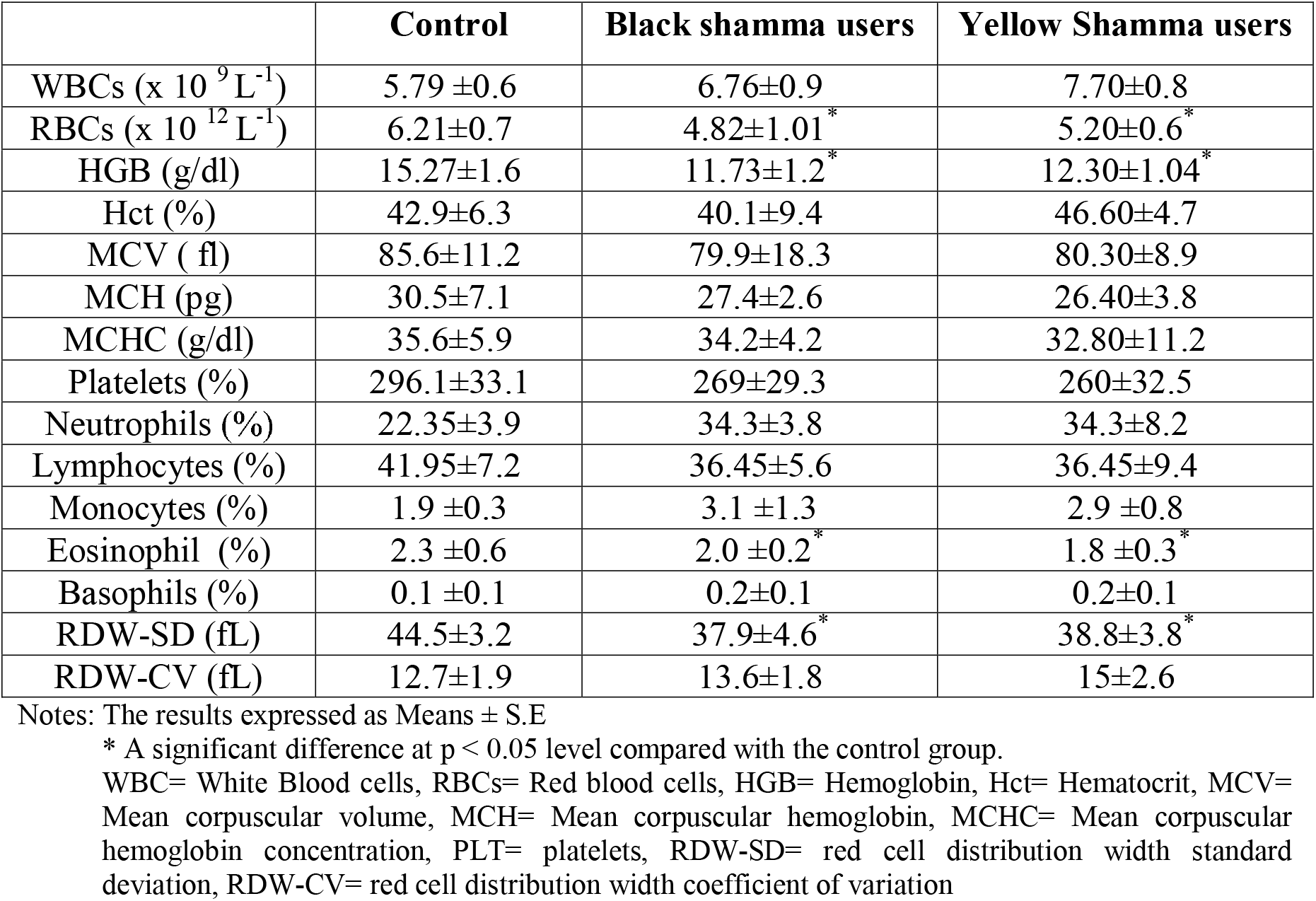
CBC analysis of normal persons not use any type of shamma (control), persons usually use black shamma and persons whom usually use yellow shamma.

|  | Control | Black shamma users | Yellow Shamma users |
| --- | --- | --- | --- |
| WBCs ($\times 10^9 \text{ L}^{-1}$) | 5.79 ± 0.6 | 6.76 ± 0.9 | 7.70 ± 0.8 |
| RBCs ($\times 10^{12} \text{ L}^{-1}$) | 6.21 ± 0.7 | 4.82 ± 1.01<sup>*</sup> | 5.20 ± 0.6<sup>*</sup> |
| HGB (g/dl) | 15.27 ± 1.6 | 11.73 ± 1.2<sup>*</sup> | 12.30 ± 1.04<sup>*</sup> |
| Hct (%) | 42.9 ± 6.3 | 40.1 ± 9.4 | 46.60 ± 4.7 |
| MCV (fl) | 85.6 ± 11.2 | 79.9 ± 18.3 | 80.30 ± 8.9 |
| MCH (pg) | 30.5 ± 7.1 | 27.4 ± 2.6 | 26.40 ± 3.8 |
| MCHC (g/dl) | 35.6 ± 5.9 | 34.2 ± 4.2 | 32.80 ± 11.2 |
| Platelets (%) | 296.1 ± 33.1 | 269 ± 29.3 | 260 ± 32.5 |
| Neutrophils (%) | 22.35 ± 3.9 | 34.3 ± 3.8 | 34.3 ± 8.2 |
| Lymphocytes (%) | 41.95 ± 7.2 | 36.45 ± 5.6 | 36.45 ± 9.4 |
| Monocytes (%) | 1.9 ± 0.3 | 3.1 ± 1.3 | 2.9 ± 0.8 |
| Eosinophil (%) | 2.3 ± 0.6 | 2.0 ± 0.2<sup>*</sup> | 1.8 ± 0.3<sup>*</sup> |
| Basophils (%) | 0.1 ± 0.1 | 0.2 ± 0.1 | 0.2 ± 0.1 |
| RDW-SD (fL) | 44.5 ± 3.2 | 37.9 ± 4.6<sup>*</sup> | 38.8 ± 3.8<sup>*</sup> |
| RDW-CV (fL) | 12.7 ± 1.9 | 13.6 ± 1.8 | 15 ± 2.6 |

Notes: The results expressed as Means ± S.E
\* A significant difference at $p < 0.05$ level compared with the control group.
WBC= White Blood cells, RBCs= Red blood cells, HGB= Hemoglobin, Hct= Hematocrit, MCV= Mean corpuscular volume, MCH= Mean corpuscular hemoglobin, MCHC= Mean corpuscular hemoglobin concentration, PLT= platelets, RDW-SD= red cell distribution width standard deviation, RDW-CV= red cell distribution width coefficient of variation

**Table 3:**
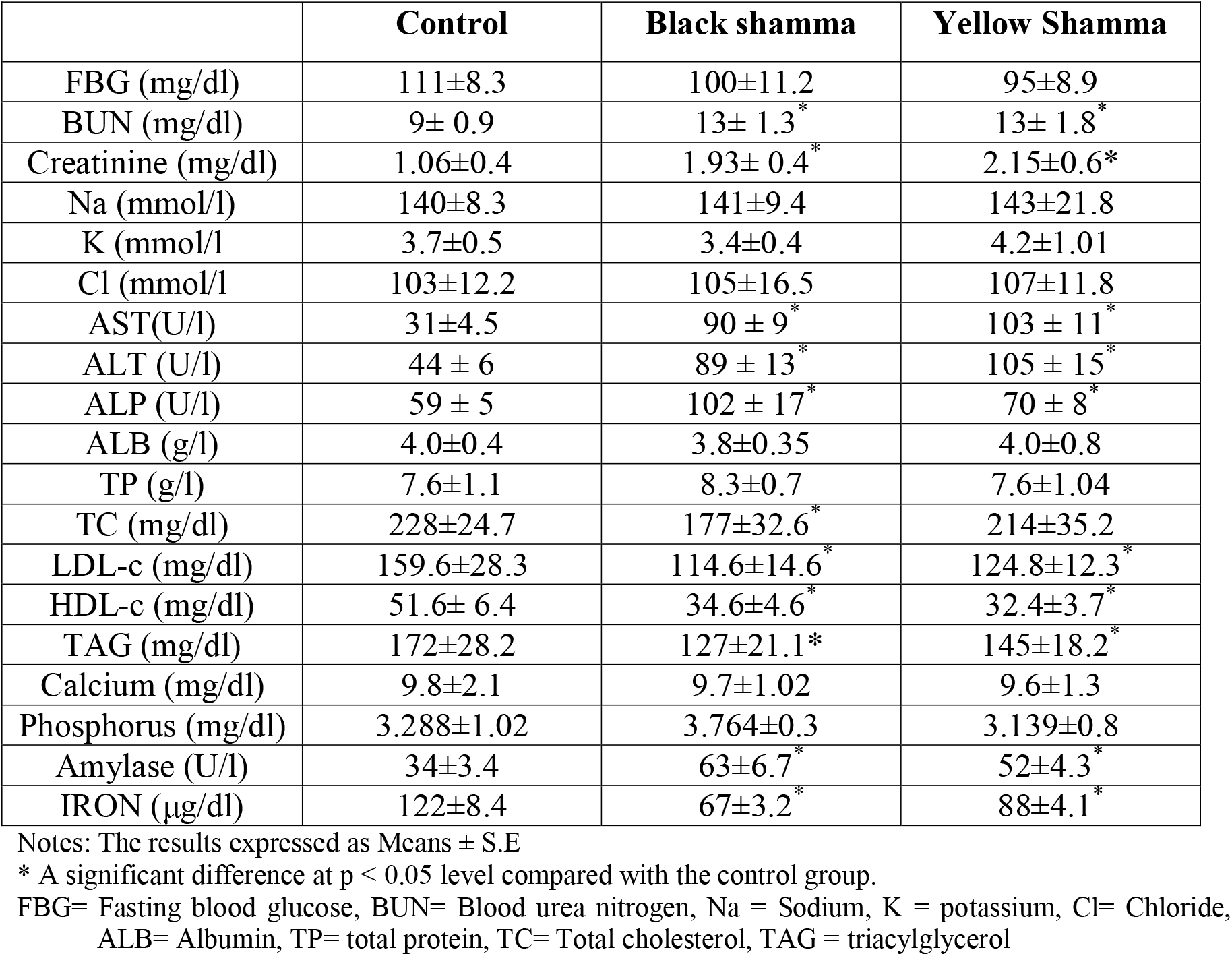
Clinical picture of normal persons not use any type of shamma (control), persons usually use black shamma and persons whom usually use yellow shamma.

The results showed in table 3 represented that, there were a significant increase in the serum levels of hepatic enzymes ALT, AST and ALP activities as well as BUN and serum creatinine concentration and amylase enzyme activity. At the same time there were a significant decrease in the serum TC, LDL-c, HDL-c, TAG and iron concentration in the same peoples; these results suggests hepatic and renal cell toxicity with lipid profile reduction and decrease in iron concentration that may be the cause of hypochromic anemia detected in table 2.

## DISCUSSION

Smokeless tobacco (shammah) is usually used by placing about 10 g into the mouth cavity between the gum and the lip, under the cheek or below the tongue on the floor of the mouth. It is then sucked slowly for a period of 15 minutes to few hours, which vary from person to person (The saliva generated meanwhile is swallowed, resulting in significantly high concentration of *shamma* components ingested. From previous studies shamma is containing a different materials with different additives, and flavors with at least 50 carcinogenic substances **[20]**. (The degree and intensity of exposure to carcinogens in terms of duration and quantity is crucial in developing cancer **[21]**. Studies regarding the constituents of *shamma* are very scarce and more emphasis has been given to the phytochemical analysis of smokeless tobacco. (so this study aimed to detect the chemicals added to smokeless tobacco during the manufacture of *shamma* and to evaluate their overall toxicity. Deleterious different effects reported about shamma as well as different types of it pushed up analyze different types of shamma present in local markets; at the same time studying its potential toxic effects on some blood parameters.

From the obtained results, the principle toxic compound detected in all shamma samples was nicotine that its predominant metabolite cotinine have been studied for its antidepressant activity. Despite the fact that nicotine itself is not carcinogenic but its derivatives such as N′-nitrosonornicotine and 4-(methylnitrosamino)-1-(3-pyridyl)-1-butanone might be responsible for the carcinogenic effect **[14]**. Other highly toxic and carcinogenic compounds were also identified in *shamma* samples such as *n*-alkanes (tridecane, tetradecane, pentadecane) (table 1) that reported to cause chemical pneumonitis lung cancer and slow death when aspired into the lungs **[22]**. It has been also reported that, alkanes may be harmful if taken in higher quantities by ingestion, inhalation skin absorption **[23]** resulting in mucous membranes irritation, enhancing mutagenesis and resulting in liver and kidney affections **[22]**.

Siloxane and its derivatives Cyclohexasiloxane and dodecamethyl have been also detected; they have been reported for their widely used as conditioning agents, emollients, in personal care products, lubricants, defoaming and antimicrobial agents **[24]**. Constituents like 3-(methylnitrosamino)-proprionitrile and nitrosamines have been reported to induce the production of reactive oxygen species in ST, leading eventually to fibroblast, DNA, and RNA damage with carcinogenic effects in the mouth of tobacco consumers. The metabolic activation of nitrosamine in tobacco by cytochrome P450 enzymes may lead to the formation of *N*-nitrosonornicotine, a major carcinogen, and micronuclei, which are an indicator of genotoxicity. (these effects further enhance DNA damage, which eventually lead to oral cancer **[25]**.

Interestingly, 3-isopropoxy-1,1,1,7,7,7-hexamethyl-3,5,5-tris(trimethylsiloxy) tetrasiloxane which is one of the components in *khat* plant (*Catha edulis*) was also identified in the shamma sample in this experiment. (It has indicated the possibility of mixing *khat* plant with *shamma* by some of the manufacturers. *Khat* is a well-known CNS stimulant with amphetamine-like properties **[26]**.

Moderate and reversible toxicity along with weight loss was found in the esophagus, stomach, liver, kidneys, and lungs **[12]**. (Presence of ammonia, benzo[a]pyrene, cadmium, nickel, nicotine, nitrates, and tobacco-specific nitrosamines increase the risk of probabilistic cancer **[27]**. Heavy metals such as arsenic and nickel have synergistic effects with risk factors associated with oral cancer **[7]**.

Biochemically, the toxicity we observed by analyzing the CBC and biochemical parameters in people usually use shamma several times represented mainly in the reduction of both RBCs count and HGB concentration which may reflects a state of hypochromic anemia; these anemia may be due to the reduction of serum iron concentration observed in table 3. The RDW-SD responsible for evaluating the range of variation of red blood cell (RBC) volume was also decreased by usual using shamma. At the same time there were also a significant eosinopenia (decrease in eosinophil) count in the cases usually use shamma with no other significant changes in any other parameters in CBC analysis.

Liver enzymes are normally found within the cells of the liver. It is well known that when the liver is injured or damaged, the liver enzymes such as ALT, AST and ALP are released into the blood **[28]**. Elevated bilirubin levels can be indicative of liver disorders or blockage of bile ducts. Increased serum AST, ALT and ALP level are important markers of liver injury, attributing to the damaged structural integrity of the liver **[29]**. ALT is primarily found in the liver, making it a more specific test for detecting liver abnormalities **[30]**. These findings indicated that shamma had a moderate and reversible effect on liver function.

It is well known that one of the primary functions of the kidneys is to remove creatinine, which is the waste product of muscle breakdown, from the bloodstream. High levels of creatinine can indicate kidney failure, which can be temporary or permanent **[31]**. Creatinine is commonly measured as an index of glomerular function **[32]**. Urea is a byproduct from protein breakdown. About 90% of urea produced is excreted through the kidney **[33]** and the blood urea nitrogen (BUN) test is also used to determine if the kidneys are successfully filtering the blood. Urea nitrogen is normal in the blood at small levels, but higher levels may indicate that the individual is experiencing kidney problems **[31]**. We observed evidence of kidney toxicity following shamma administration, with increased BUN and serum creatinine levels.

From all of the above, it can be concluded that, shamma contains different concentrations of nicotine and other toxic substances and can produce its toxic effects to liver and kidney tissues as well as affecting CBC picture resulting in hypochromic anemia with iron deficiency and increasing amylase activities. So, aawareness campaigns to all people around the dangerous effect of frequent use of shamma and other types of smokeless tobacco should be regulated.

## Data Availability

Available on requiest

